# Rapid Detection of SARS-CoV-2 Antigen from Serum in a Hospitalized Population

**DOI:** 10.1101/2020.12.21.20248140

**Authors:** K McAulay, EJ Kaleta, TE Grys

## Abstract

SARS-CoV-2 viremia has been demonstrated in some patients using molecular assays. Here we demonstrate detection of SARS-CoV-2 antigen in a cohort of hospitalized patients using a rapid diagnostic test from Anhui Deepblue Medical Technology Co., Ltd. We detected antigen in serum from 11 of 13 patients at time points ranging from three to eighteen days from symptom onset and observed that the disappearance of an antigen signal was associated with seroconversion. These results demonstrate proof of principle use of a rapid antigen test with serum samples in a format compatible with point of care testing.

## Introduction

The coronavirus disease 2019 (COVID-19) pandemic, caused by severe acute respiratory syndrome virus 2 (SARS-CoV-2), has placed tremendous strain on centralized clinical laboratories due to high volumes of testing and supply chain challenges. Rapid diagnostic tests (RDT) present an opportunity to reduce the burden on centralized testing labs. Many RDT are lateral flow devices which, with appropriate validation, can easily be performed in a point of care (POC) setting. There are a large number of these tests currently approved by the FDA for antibody detection and the market for rapid antigen detection from respiratory specimens is currently expanding. While concerns remain regarding clinical accuracy[1, 2], these tests are changing the response to the pandemic since the results can be available in as little as 10-20 minutes after specimen collection. This compares favorably to the current gold standard molecular methods, for which laboratory turn-around times for high-throughput batch testing are typically 6-8 hours or more, and when there is a surge of disease or shortage of supplies, actual turn-around may take days.

SARS-CoV-2 is known to reach high levels of detectable RNA in the nasopharynx. While there have been several reports of measurable viremia in hospitalized patients, these investigations have focused on nucleic acid detection[3, 4]; there are few peer-reviewed reports of SARS-CoV-2 antigenemia[5, 6]. Since some viral proteins are present in multiple copies per virion, we tested whether a rapid antigen kit could detect antigenemia in patients who had been diagnosed with COVID-19.

## Methods

### Clinical Specimens

Residual serum was obtained from 54 standard of care specimens from 13 SARS-CoV-2 PCR-positive, hospitalized patients at Mayo Clinic Arizona and stored at - 80 °C until further use. The serum reflected samples collected from 2 to 24 days after onset of symptoms. Use of these specimens was reviewed and approved by the Mayo Clinic Institutional Review Board under protocol # 20-004544.

### Antibody Tests

Antibody tests were performed using the SARS-CoV-2 IgG/IgM Rapid Test (ACON Laboratories Inc.) according to the manufacturer’s instructions. Briefly, 5 μL serum was applied to the sample well, followed by 2-3 drops of buffer. Tests were incubated on a flat surface and read after 15 minutes. Results were recorded in a binary fashion as either positive or negative, with IgM alone, IgG alone, or IgM + IgG all being recorded as “positive”.

### Antigen Tests

Antigen tests were performed using COVID-19 (SARS-CoV-2) Antigen Test Kit (Colloidal Gold) (Anhui Deepblue Medical Technology Co., Ltd.) using the same process described above for antibody tests. i.e. 5 μL serum was applied to the sample well, followed by 2-3 drops of buffer. Tests were incubated on a flat surface and read after 15 minutes.

## Results & Discussion

Serum from 13 hospitalized SARS-CoV-2 RT-PCR-positive patients at Mayo Clinic Arizona were tested for SARS-CoV-2 antigen and antibody using COVID-19 (SARS-CoV-2) Antigen Test Kit (Colloidal Gold) (Anhui Deepblue Medical Technology Co., Ltd.) and SARS-CoV-2 IgG/IgM Rapid Test (ACON Laboratories Inc.), respectively. Specimens spanned 2-24 days from the onset of symptoms with a minimum of two and a maximum of eight time points per patient (n = 54). For 12 of the 13 patients tested (51/54 specimens), serum was always positive for either antigen, antibody, or both, and disappearance of antigen was always associated with the presence of antibody. The latest antigen detection occurred 18 days after symptom onset and the earliest antibody detection occurred six days after symptom onset.

The potential to offer serology testing in a POC setting, such as a drive-through, provides an opportunity to avoid centralized phlebotomy, thus minimizing the risk of exposure for patients. However, what is particularly striking here is detection of SARS-CoV-2 antigen in the serum of 11 of 13 individuals using an assay designed for pharyngeal and nasopharyngeal specimens. While a small number of studies have described detectable SARS-CoV-2 antigenemia [5,6], to our knowledge this is the first such report using rapid tests with the potential to be used in a POC setting. The authors recognize that these findings must be validated using a larger sample size and including non-hospitalized individuals with mild or asymptomatic infection to verify the sensitivity of antigen detection in serum. Nonetheless, an opportunity to rapidly detect COVID-19 infection status at the POC, without respiratory specimen collection, is promising. Further, detectable antigenemia may be a useful prognostic indicator for disease acuity in hospitalized patients and may potentially be used to demonstrate response to therapies; this novel application therefore warrants further study.

**Table 1.**
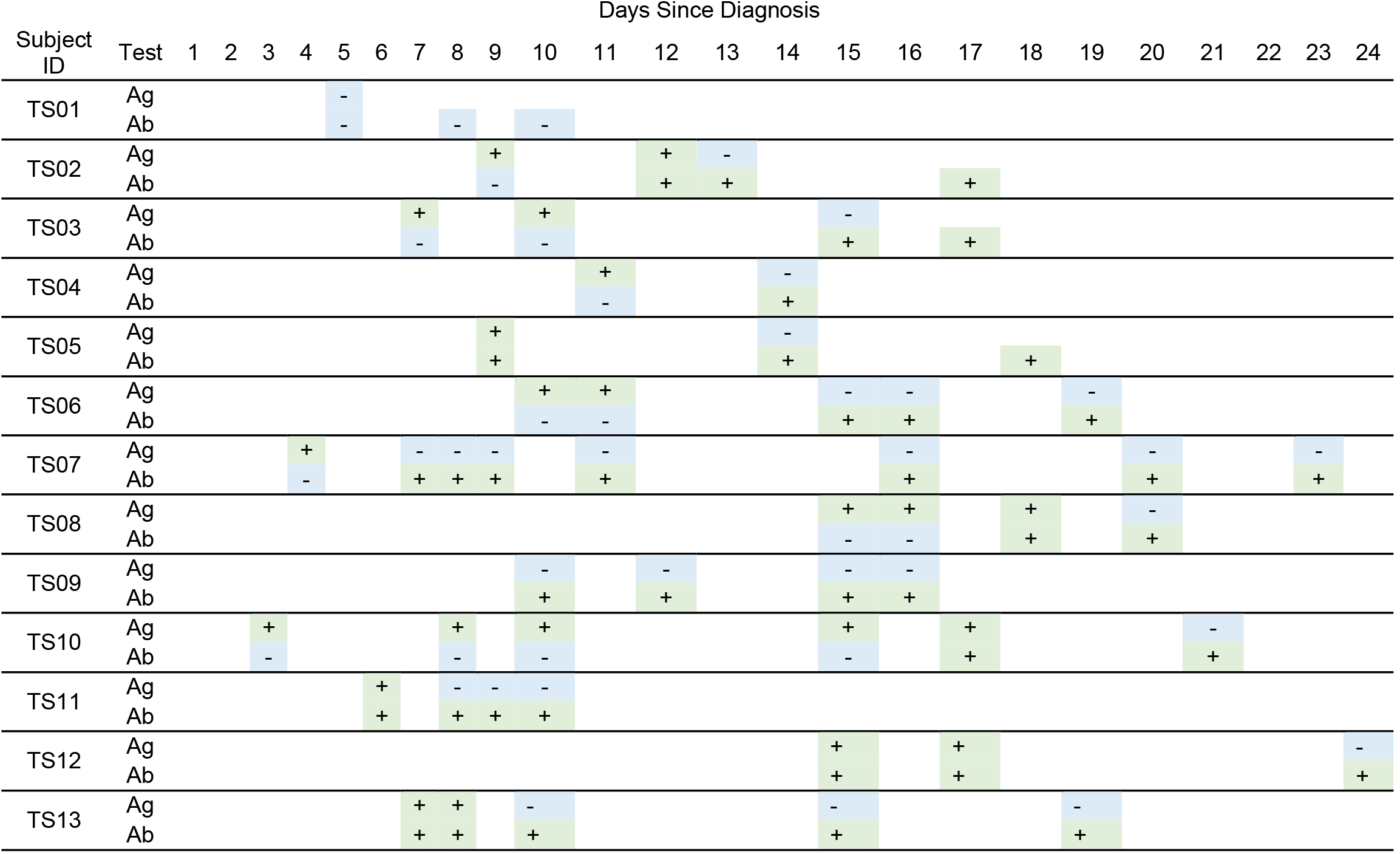
Serum antigen (Ag) and antibody (Ab) results for 13 SARS-CoV-2 PCR-positive, hospitalized individuals as determined by COVID-19 (SARS-CoV-2) Antigen Test Kit (Colloidal Gold) (Anhui Deepblue Medical Technology Co., Ltd.) and SARS-CoV-2 IgG/IgM Rapid Test (ACON Laboratories Inc.), respectively.

**Figure 1.**
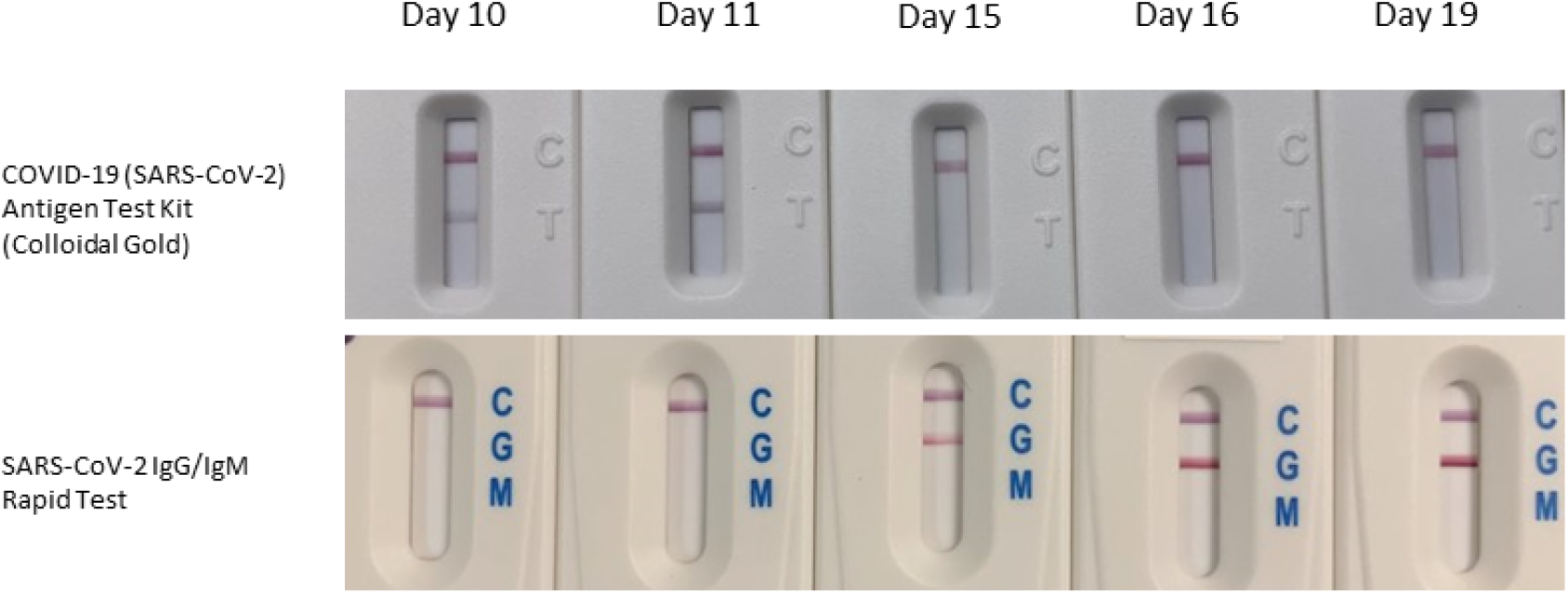
Serum antigen and antibody results for subject TS06 determined by COVID-19 (SARS-CoV-2) Antigen Test Kit (Colloidal Gold) (Anhui Deepblue Medical Technology Co., Ltd.) and SARS-CoV-2 IgG/IgM Rapid Test (ACON Laboratories Inc.), respectively at 10, 11, 15, 16, and 19 days after symptom onset.

## Data Availability

All data generated for this brief proof of concept study is included in this article.

## Acknowledgements

The authors wish to thank Paramount Labs for providing the Anhui DeepBlue kits and ACON Laboratories for providing the antibody kits. We also recognize salary support for KM and TG and from the Mayo Clinic Center for Individualized Medicine.

